# A Multi-Omics Computational Pipeline for Systematic Discovery of Retired Self-Antigens as Cancer Vaccine Targets

**DOI:** 10.64898/2026.04.20.26351288

**Authors:** V Wang, S Deng, R Aguilar

**Author notes:** Corresponding author: Roberto Aguilar |. Dedicated to the memory of Vincent K. Tuohy, Ph.D. (1947–2024), whose vision of cancer immunoprevention inspired this work.

## Abstract

**Background:** The retired antigen hypothesis, introduced by Tuohy and colleagues, proposes that tissue-specific proteins expressed conditionally during early life or reproductive stages, then silenced in normal aging tissue, represent safe and effective cancer vaccine targets when re-expressed in tumors. To date, discovery of retired antigens has relied entirely on hypothesis-driven wet lab work, limiting throughput.

**Methods:** Here we present RADAR (Retired Antigen Discovery and Ranking), a multi-omics computational pipeline implemented on a standard server that systematically identifies retired antigen candidates. RADAR comprises four core discovery layers integrating: 1) The Genotype-Tissue Expression Portal (GTEx) normal tissue expression, 2) TCGA tumor re-expression, 3) DNA methylation, and 4) miRNA regulatory networks, each applied sequentially to identify genes exhibiting the epigenetic and post-transcriptional hallmarks of tissue-specific retirement followed by tumor re-activation. Candidate characterization is further supported by three automated modules: 1) protein-level safety screening via the Human Protein Atlas, 2) molecular subtype enrichment analysis, and 3) cross-cancer confirmation, which execute automatically when the relevant data are available for the selected cancer type.

**Results:** The pipeline independently validated known targets including alpha-lactalbumin (LALBA, the basis of the Tuohy Phase 1 triple-negative breast cancer vaccine trial) and anti-Mullerian hormone (AMH), consistent with Tuohy’s ovarian cancer vaccine program targeting AMHR2, and rediscovered multiple known cancer-testis antigens (MAGEA1, MAGEC1, SSX1) as positive controls. Among 4,664 initial candidates derived from GTEx, the pipeline identified 20 high-confidence retired antigen candidates passing all filters. DCAF4L2, COX7B2, TEX19, and CT83 emerge as the highest-priority novel candidates for experimental validation, demonstrating zero expression in critical somatic organs, strong epigenetic silencing, and significant re-expression across multiple cancer types.

**Conclusion:** RADAR provides the first systematic computational framework for retired antigen discovery, offering a reproducible and scalable approach to expanding the cancer immunoprevention pipeline beyond individually characterized targets. The pipeline is fully reproducible, requires no specialized hardware, and is immediately extensible to additional TCGA cancer types.

## 1. Introduction

Cancer vaccines targeting tumor-associated antigens represent a promising but largely unrealized strategy for cancer prevention and treatment. The central challenge is identifying antigens that are sufficiently tumor-specific to elicit a therapeutic immune response without inducing autoimmune damage to normal tissues (Fan *et al*., 2023). Neoantigens arising from somatic mutations offer high specificity but are patient-unique, limiting their utility for population-level prevention strategies (Xie *et al*., 2023).

Tuohy and colleagues proposed an elegant alternative: the retired antigen hypothesis. Self-proteins expressed conditionally during specific life stages such as lactation, reproductive activity, or early development, and subsequently silenced in normal aging tissues, represent a distinct class of cancer vaccine targets (Tuohy *et al*., 2020; Tuohy, 2014). When these proteins are re-expressed in tumors, the immune system has effectively lost tolerance to them due to their prolonged absence, enabling a vaccine-induced response (Tuohy *et al*., 2020; Tuohy, 2014). Critically, because the proteins are not expressed in normal adult somatic tissues, autoimmune risk is substantially lower than for conventional tumor-associated antigens.

Alpha-lactalbumin (LALBA), a milk protein expressed only in lactating breast tissue, exemplifies this concept. Tuohy and colleagues demonstrated that LALBA is re-expressed in triple-negative breast cancer (TNBC), and a vaccine targeting LALBA completed Phase 1 clinical trials at Cleveland Clinic, demonstrating safety and protocol-defined immune responses in 74% of participants (Jaini *et al*., 2010; Johnson *et al*., 2025). Phase 2 trials are now being planned. Similarly, the extracellular domain of anti-Mullerian hormone receptor II (AMHR2-ED), expressed in reproductive-stage ovarian tissue, was identified as a target for ovarian cancer prevention, with preclinical studies demonstrating significant tumor inhibition (Mazumder et al., 2017; Sakalar *et al*., 2015).

Despite these successes, discovery of retired antigen candidates has relied entirely on hypothesis-driven wet lab work, limiting throughput and scalability. No systematic computational framework for retired antigen discovery has been reported. The authors of the present study were directly involved in the preclinical development of retired antigen vaccine strategies for breast, ovarian, and testicular cancers (Aguilar *et al*., 2017; Sakalar *et al*., 2015; Mazumder *et al*., 2017), providing the biological foundation and motivation for developing a systematic computational discovery framework. Furthermore, the late Vincent K. Tuohy, Ph.D., whose foundational work established the retired antigen hypothesis and brought it to clinical application, passed away before the full potential of this approach could be realized computationally. Extending his vision through systematic computational discovery represents both a scientific opportunity and a tribute to his enduring contribution to cancer immunoprevention.

Here we present RADAR (Retired Antigen Discovery and Ranking), a multi-omics computational pipeline that integrates publicly available databases including The Genotype-Tissue Expression Portal (GTEx), the Human Protein Atlas (HPA), and TargetScan with institutional TCGA multi-omics data to screen 56,200 genes through four sequential discovery layers. RADAR produces a ranked priority list of novel retired antigen candidates, further characterized by three automated modules assessing protein-level safety, molecular subtype enrichment, and cross-cancer confirmation. We demonstrate RADAR using breast and ovarian cancer as validation examples, independently confirming known targets and identifying novel high-priority candidates for experimental validation.

## 2. Methods

### 2.1 Computational Infrastructure

All analyses were performed on a standard server running Ubuntu Server 24.04 LTS, equipped with a 6-core Intel Xeon processor, 32 GB RAM, and approximately 20 TB of storage. No GPU acceleration was required. All Python analyses were performed within a conda environment using Python 3.10, pandas, numpy, and pyarrow, demonstrating that RADAR is deployable on hardware commonly available in academic research settings without specialized computational infrastructure.

### 2.2 Data Sources

Four primary data sources support the four core discovery layers, with three supplementary sources used by the automated characterization modules:

#### Primary Data Sources

- GTEx v8 (August 2019): Gene median transcripts per million (TPM) across 54 tissues and 56,200 genes, downloaded from the GTEx portal (adult-gtex Google Cloud bucket). Subject phenotype files including age brackets spanning 20-79 years were retained for future age-stratified analyses. Used by Layer 1 (tissue-specific silencing).
- TCGA multi-omics expression data: Pre-computed differential expression results across 15 cohorts including 14 TCGA cancer types: bladder urothelial carcinoma (BLCA), breast invasive carcinoma (BRCA), cervical squamous cell carcinoma and endocervical adenocarcinoma (CESC), colon adenocarcinoma (COAD), glioblastoma multiforme (GBM), kidney renal clear cell carcinoma (KIRC), liver hepatocellular carcinoma (LIHC), lung adenocarcinoma (LUAD), pancreatic adenocarcinoma (PAAD), prostate adenocarcinoma (PRAD), skin cutaneous melanoma (SKCM), stomach adenocarcinoma (STAD), thyroid carcinoma (THCA), and uterine corpus endometrial carcinoma (UCEC), and one TARGET pediatric cohort (TARGET-ALL-P3), stored as parquet files (969,856 entries). Used by Layer 2 (tumor re-expression).
- DNA methylation: Pan-cancer methylation M-values across 360,872 CpG probes for cancer and matched normal tissue, stored as parquet files on an institutional server. Used by Layer 3 (epigenetic silencing).
- miRNA and TargetScan v8: Pan-cancer differential expression results for 1,881 miRNAs across 12 TCGA cohorts, combined with predicted miRNA target sites from TargetScan v8 (human, species ID 9606). Used by Layer 4 (post-transcriptional silencing).

#### Supplementary Data Sources

- Human Protein Atlas (downloaded November 2023): Protein expression, subcellular localization, tissue specificity, and TCGA-derived cancer prognostic data for 20,162 genes, accessed via proteinatlas.tsv. Used by the protein-level safety screening module.
- TCGA-OV (ovarian serous cystadenocarcinoma): 20 primary tumor gene expression files downloaded from the GDC API (STAR-Counts workflow, TPM values), compared against GTEx ovary tissue as the normal baseline. Used as an example application of the cross-cancer confirmation module.
- TCGA BRCA subtype annotations: Molecular subtype classifications (Luminal A, Luminal B, Basal-like, HER2-enriched) obtained from the UCSC Xena BRCA clinical matrix. TNBC status was defined as ER-negative, PR-negative, and HER2-negative by IHC (n=130 TNBC, n=1,051 non-TNBC). Used as an example application of the subtype enrichment module.

### 2.3 Pipeline Layers

#### Layer 1: GTEx Tissue-Specific Silencing

All 56,200 genes in GTEx (GTEx Consortium, 2020) were screened for tissue-specific expression in reproductive tissues (breast mammary tissue, ovary, uterus, fallopian tube, vagina, prostate, testis, and cervix uteri) and low expression in adult normal somatic tissues (18 tissues including heart, brain, liver, kidney, lung, colon, skin, muscle, and whole blood). Candidates were required to have a maximum reproductive tissue TPM greater than 5 and a mean adult somatic tissue TPM less than 2. The lower bound of TPM greater than 5 in reproductive tissue was chosen to ensure meaningful expression above background noise while remaining inclusive of genes with moderate tissue-specific expression. The upper bound of mean TPM less than 2 in adult somatic tissues was chosen to exclude genes with appreciable expression in non-reproductive tissues, minimizing autoimmunity risk while retaining genes with low but detectable somatic expression that may reflect residual or tissue-specific isoform expression. This yielded 4,664 initial candidates.

#### Layer 2: TCGA Tumor Re-expression

Candidates from Layer 1 were cross-referenced against the pan-cancer differential expression dataset derived from TCGA (Tomczak *et al*., 2015). Genes significantly upregulated (log2FC > 1) in tumor versus normal tissue in at least one TCGA cohort were retained. This filter confirms the essential second criterion of the retired antigen hypothesis: that a gene silenced in normal adult tissue must be re-expressed in tumor tissue to present a targetable antigen. A threshold of log2FC greater than 1, representing a minimum two-fold increase in expression, was applied as a conventional cutoff for biologically meaningful differential expression in RNA-seq analyses (Love et al., 2014). Requiring significance in at least one cohort rather than across all cohorts reflects the cancer-type specificity of retired antigen re-expression, whereby a candidate may be relevant as a vaccine target in one or more cancer types without being universally re-expressed. This reduced the candidate list to 1,729 genes distributed across up to 15 cancer types.

#### Layer 3: DNA Methylation Epigenetic Silencing

The M-value represents the log2 ratio of methylated to unmethylated signal intensity at each CpG probe, where positive values indicate methylation and negative values indicate unmethylation (Du *et al*., 2010). Candidate genes were screened for the epigenetic retirement signature, defined as high promoter methylation in normal tissue (maximum probe M-value > 2.5) with hypomethylation in tumor tissue (delta M-value < -0.3). This signature confirms that the tissue-specific silencing identified in Layer 1 is mediated by DNA methylation, providing mechanistic evidence that the gene has been epigenetically retired in normal adult tissue. Genes meeting this criterion are more likely to exhibit stable, heritable silencing that is progressively lost during carcinogenesis, a defining characteristic of retired antigen biology (Baylin and Jones, 2016). M-values were computed per gene by averaging across all associated CpG probes. This filter reduced candidates from 1,729 to 84, confirming that epigenetic silencing is the primary mechanism of tissue-specific retirement and that only a small fraction of tissue-specific genes achieve their specificity through true epigenetic retirement.

#### Layer 4: miRNA Regulatory Network

MicroRNAs (miRNAs) are small non-coding RNAs that suppress gene expression post-transcriptionally by binding to the 3’ untranslated region of target messenger RNAs, leading to their degradation or translational repression (Bartel, 2004). In the context of retired antigen biology, miRNA-mediated silencing represents a second independent mechanism by which tissue-specific genes may be suppressed in normal adult tissue and de-repressed in tumor tissue. When miRNAs that target a candidate gene are themselves downregulated in tumor tissue, the candidate gene loses a key post-transcriptional brake on its expression, contributing to its re-activation in cancer (Esquela-Kerscher and Slack, 2006; Garzon *et al*., 2009). Requiring evidence of miRNA-mediated regulation therefore provides orthogonal mechanistic support for the retirement signature identified in Layers 1 through 3.

MiRNAs significantly downregulated in tumor versus normal tissue (log2FC < -1) across available TCGA cohorts were extracted from the pan-cancer miRNA dataset. Predicted target genes for these miRNAs were identified using TargetScan v8 (Agarwal *et al*., 2015), which provides computationally predicted miRNA binding sites based on seed sequence complementarity and evolutionary conservation. Candidate genes targeted by two or more downregulated miRNAs were retained, as convergent regulation by multiple miRNAs provides stronger evidence of coordinated post-transcriptional silencing than regulation by a single miRNA alone (Krek *et al*., 2005). This approach identified a co-regulated post-transcriptional silencing network contributing to tissue-specific retirement across cancer types.

### 2.4 Characterization Modules

The following three modules execute automatically when the relevant data are available for the selected cancer type. They do not filter candidates but characterize and prioritize them across additional biological dimensions.

#### Module 1: Protein-Level Safety Screening

Candidates were cross-referenced against the Human Protein Atlas for protein-level tissue specificity confirmation, subcellular localization to determine surface accessibility for antibody-based vaccine strategies, and TCGA-derived cancer prognostic data. Autoimmunity safety screening was performed using GTEx critical organ expression across heart, brain, liver, kidney, lung, blood, muscle, colon, stomach, and skin. Candidates with maximum TPM greater than 5 in any critical organ were flagged as autoimmunity risks and deprioritized.

#### Module 2: Molecular Subtype Enrichment

When molecular subtype annotations are available for the selected cancer type, candidates are evaluated for enrichment within clinically relevant subtypes. As a demonstration, TCGA BRCA samples were stratified by molecular subtype using UCSC Xena annotations. The 130 confirmed TNBC samples (ER-negative, PR-negative, HER2-negative by IHC) were compared against 1,051 non-TNBC samples using average log2(TPM+1) values. Candidates showing enrichment in TNBC (delta log2TPM > 0.3) were scored accordingly.

#### Module 3: Cross-Cancer Confirmation

When expression data are available for a second related cancer type, candidates are evaluated for re-expression in that cancer as additional evidence of broad retired antigen biology. As a demonstration, average TPM across 20 TCGA-OV primary tumor samples was compared against GTEx ovary tissue TPM as the normal baseline. Log2 fold change was computed as log2((mean tumor TPM + 1) / (GTEx ovary TPM + 1)). Candidates upregulated in both the primary and secondary cancer types were flagged as dual-cancer targets.

### 2.5 Composite Scoring

A composite score was computed for each candidate gene integrating all pipeline layers, adapted from the multi-omics scoring framework described by Chen *et al*. (2020). The final score was calculated as:

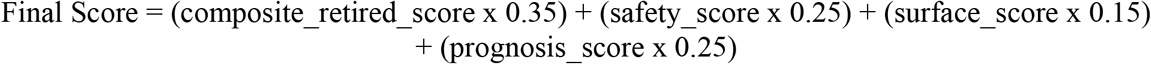

Where composite_retired_score = (max_log2FC x 0.4) + (|delta methylation| x 0.3) + (n_cancers x 0.3), with max_log2FC representing the maximum log2 fold change in tumor versus normal tissue across all available TCGA cohorts, and n_cancers representing the number of TCGA cohorts in which the gene is significantly upregulated. Safety_score was assigned as 2.0 if maximum TPM in critical organs was less than 1.0, 1.0 if less than 5.0, and 0.0 otherwise, reflecting the prioritization of candidates with the lowest autoimmunity risk. Surface_score was assigned as 1.0 if the candidate was surface-accessible per HPA subcellular localization data and 0.5 otherwise. Prognosis_score was assigned as 1.0 if the candidate was associated with unfavorable prognosis in HPA TCGA-derived data, 0.5 if unprognostic, and 0.0 if favorable. Weights were assigned to reflect the relative importance of each component, with the core retired antigen signal weighted most heavily, followed by clinical relevance through safety and prognosis, and surface accessibility weighted as a secondary consideration for vaccine strategy rather than candidate prioritization.

## 3. Results

### 3.1 Pipeline Validation

To assess pipeline performance, we examined whether RADAR could independently recover known retired antigen targets and established cancer-testis antigens without prior knowledge of their identity.

Alpha-lactalbumin (LALBA), the target of the Tuohy Phase 1 TNBC vaccine trial (Tuohy *et al*., 2016), was confirmed as a retired antigen across all four core discovery layers. In GTEx, LALBA showed reproductive tissue-specific expression (breast mammary tissue TPM = 3.14) with zero expression in all critical somatic organs, satisfying the tissue-specific silencing criterion of Layer 1. Strong epigenetic silencing was confirmed in normal tissue (DNA methylation M-value = 3.14) with tumor hypomethylation (delta M-value = -0.514), satisfying Layer 3. LALBA was upregulated in TNBC tumor tissue relative to non-TNBC (delta log2TPM = +0.759), and HPA tissue specificity was classified as High. LALBA ranked 17th in the final priority list, appropriately positioned below candidates with stronger tumor re-expression signals, reflecting the pipeline’s ability to correctly weight the magnitude of tumor re-expression alongside epigenetic retirement.

AMH, the basis of Tuohy’s ovarian cancer vaccine program targeting the extracellular domain of its receptor AMHR2 (Mazumder *et al*., 2017), was correctly handled by the pipeline in two ways. First, AMH was excluded from TNBC subtype enrichment (delta log2TPM = -0.072), correctly identifying it as not enriched in breast cancer subtypes. Second, AMH was confirmed upregulated in TCGA-OV primary tumors versus GTEx ovary tissue as the normal baseline (log2FC = +1.474, mean tumor TPM = 5.847 versus GTEx ovary TPM = 1.465), demonstrating that the cross-cancer confirmation module correctly identifies ovarian cancer retired antigen candidates.

Three known cancer-testis antigens (CTAs) were independently rediscovered by the pipeline without prior knowledge of their CTA status: MAGEA1 (rank 1, final score 2.694), MAGEC1 (rank 4, final score 1.990), and SSX1 (rank 3, final score 2.073). CTAs are a well-characterized class of tumor antigens defined by testis-restricted expression in normal adult tissue and aberrant re-expression across multiple cancer types (Simpson *et al*., 2005), making them the most established biological archetype of the retired antigen concept. Their independent rediscovery by an unbiased computational pipeline confirms that RADAR’s four core discovery layers correctly capture the biological signature of tissue-specific retirement followed by tumor re-activation.

### 3.2 Pipeline Funnel

The RADAR pipeline progressively refined 56,200 screened genes to 61 high-confidence retired antigen candidates through four sequential core discovery layers, as summarized in Table 1. Following characterization by the three automated modules, 20 candidates were confirmed as high-confidence retired antigen targets after protein-level safety screening, with 10 candidates deprioritized due to expression in critical somatic organs exceeding the autoimmunity safety threshold.

**Table 1:**
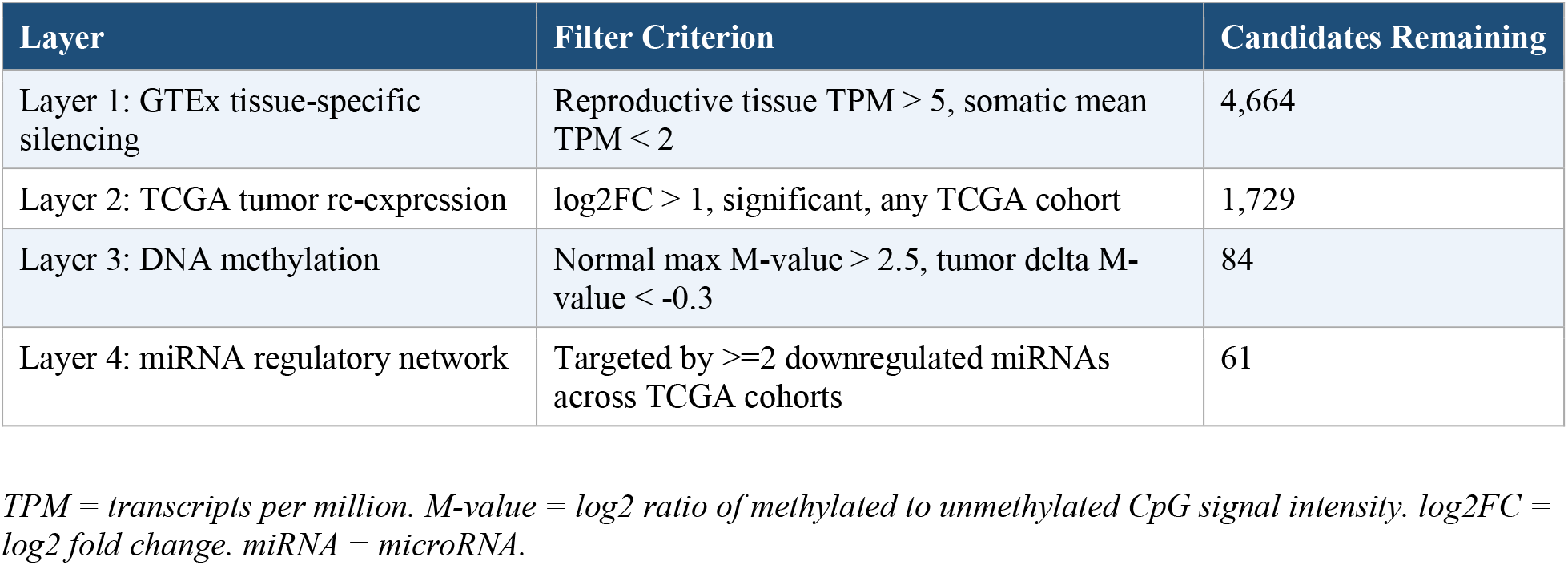
RADAR progressively refined 56,200 screened genes to 61 high-confidence retired antigen candidates through four sequential discovery layers. GTEx v8 was used to identify genes exhibiting tissue-specific expression in reproductive tissues with near-absent expression in adult somatic tissues. Candidates were subsequently filtered for significant tumor re-expression across TCGA cohorts, epigenetic silencing confirmed by DNA methylation M-values, and post-transcriptional regulation by tumor-downregulated miRNAs identified using TargetScan v8. Each layer applied independent biological criteria, and only genes satisfying all four filters were retained as high-confidence retired antigen candidates for downstream characterization.

Following core discovery, the 61 high-confidence candidates were evaluated by three automated characterization modules, as summarized in Table 2. Protein-level safety screening via the Human Protein Atlas identified 10 candidates with expression in critical somatic organs exceeding the autoimmunity safety threshold, which were subsequently deprioritized. As example applications of the subtype enrichment and cross-cancer confirmation modules, 24 candidates showed enrichment in TNBC relative to non-TNBC breast cancer subtypes, and 20 candidates were confirmed upregulated in TCGA-OV primary tumors relative to GTEx ovary tissue as the normal baseline, identifying a subset of dual-cancer retired antigen candidates with potential relevance across both breast and ovarian cancer.

**Table 2:** Three automated characterization modules evaluated and prioritized the 61 core pipeline candidates across protein-level safety, molecular subtype enrichment, and cross-cancer confirmation. Candidates passing all four core discovery layers were subsequently characterized by three automated modules that execute when relevant data are available for the selected cancer type. Module 1 screened all 61 candidates against Human Protein Atlas protein expression data in critical somatic organs, flagging candidates with maximum TPM greater than 5 as autoimmunity risks. Modules 2 and 3 are presented here as example applications using breast and ovarian cancer data.

| Module | Application | Candidates Characterized |
| --- | --- | --- |
| Module 1: Protein-level safety screening | HPA critical organ TPM < 5 | 61 evaluated, 10 flagged as autoimmunity risk |
| Module 2: Subtype enrichment | TNBC vs non-TNBC enrichment (BRCA example) | 24 enriched in TNBC |
| Module 3: Cross-cancer confirmation | TCGA-OV vs GTEx ovary (OV example) | 20 confirmed in both BRCA and OV |

*TNBC = triple-negative breast cancer. TCGA-OV = ovarian serous cystadenocarcinoma cohort. HPA = Human Protein Atlas. TPM = transcripts per million.*

### 3.3 Top Candidates

The 20 high-confidence retired antigen candidates remaining after all core discovery layers and characterization modules were ranked by integrated composite score, as presented in Table 3. Candidates are ranked in descending order of priority, with Rank 1 representing the highest composite score and therefore the highest priority for experimental validation. Known cancer-testis antigens MAGEA1, MAGEC1, and SSX1 ranked among the top four candidates, validating pipeline specificity through independent rediscovery of established retired antigen archetypes. The known retired antigen LALBA ranked 17th, appropriately reflecting its moderate tumor re-expression signal relative to novel candidates. Among novel protein-coding candidates, DCAF4L2, COX7B2, TEX19, and CT83 emerged as the highest priority targets for experimental validation, demonstrating strong epigenetic silencing in normal tissue, significant tumor re-expression across multiple cancer types, and zero expression in critical somatic organs. Surface-accessible candidates suitable for antibody-based vaccine strategies included COL22A1, HHIPL2, CPA6, and LALBA, while the remaining candidates are intracellular and would require peptide, mRNA, or viral vector vaccine formulations to elicit T-cell mediated responses.

**Table 3:**
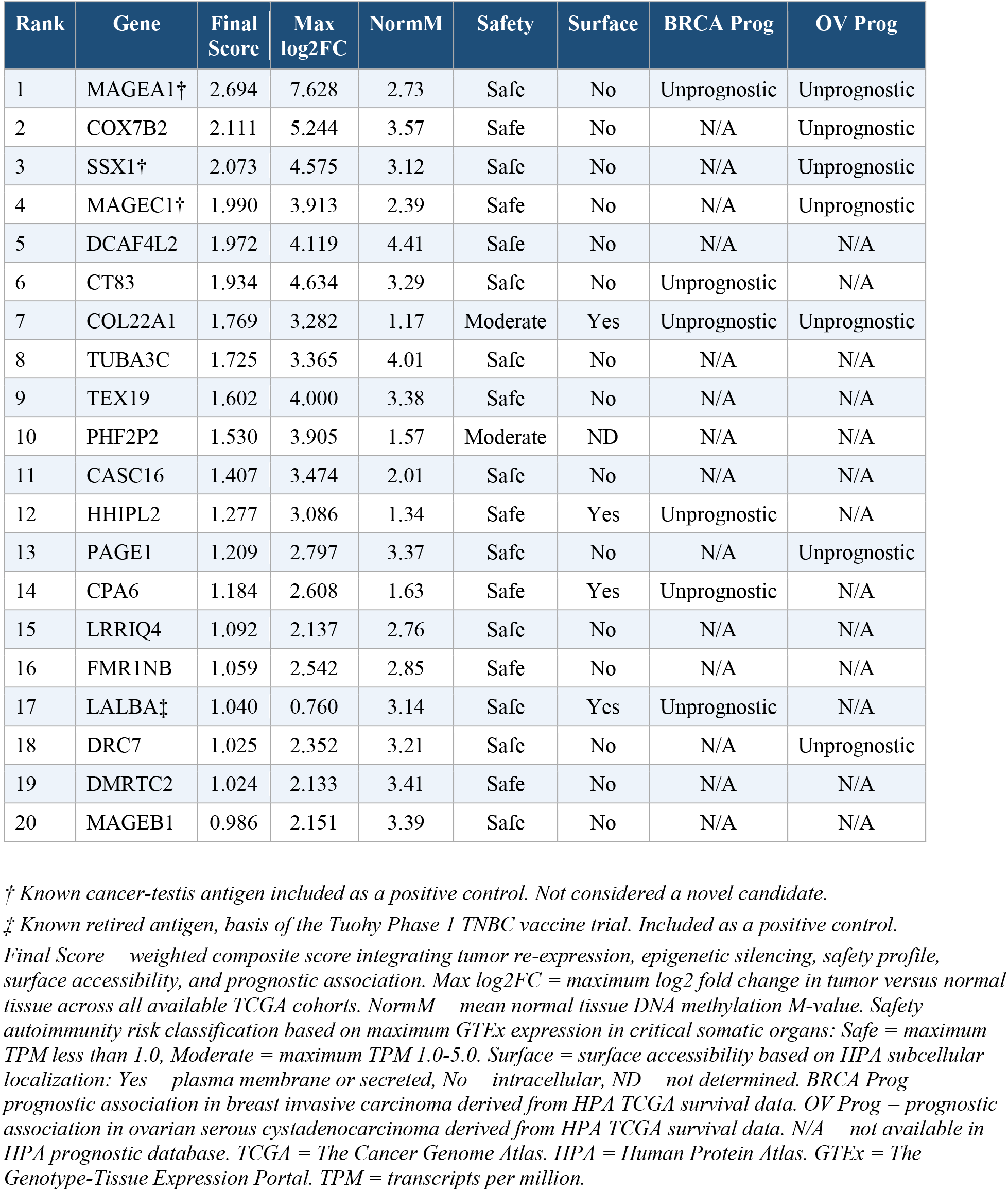
Twenty high-confidence retired antigen candidates ranked by integrated composite score following all core discovery layers and characterization modules. Candidates were scored using a weighted composite formula integrating the core retired antigen signal, autoimmunity safety profile, surface accessibility, and prognostic association. Known cancer-testis antigens and known retired antigens are marked and included as positive controls to validate pipeline specificity.

### 3.4 High-Priority Candidates

Given that MAGEA1, MAGEC1, and SSX1 are established cancer-testis antigens discussed in section 3.1, the following candidates represent the highest-priority targets identified by RADAR, comprising both novel candidates and underexplored known cancer-testis antigens.

#### COX7B2 (Rank 2)

Cytochrome c oxidase subunit 7B2 (COX7B2) is a mitochondrial respiratory chain component whose expression in GTEx is restricted to testis, consistent with the cancer-testis antigen expression pattern (GTEx Consortium, 2020). COX7B2 demonstrates strong epigenetic retirement in normal tissue (normal M-value = 3.57) and is significantly upregulated across 6 cancer types including bladder urothelial carcinoma, breast invasive carcinoma, liver hepatocellular carcinoma, lung adenocarcinoma, prostate adenocarcinoma, and stomach adenocarcinoma (max log2FC = 5.244). It shows zero expression in all critical somatic organs (max TPM = 0.000), indicating a favorable autoimmunity safety profile. COX7B2 has not previously been characterized as a retired antigen vaccine candidate and represents a novel high-priority target for experimental validation.

#### DCAF4L2 (Rank 5)

DDB1 and CUL4 associated factor 4 like 2 (DCAF4L2) shows the strongest normal tissue methylation signal in the entire candidate list (normal M-value = 4.41), indicating deep epigenetic retirement. It is significantly upregulated across 6 cancer types including bladder urothelial carcinoma, breast invasive carcinoma, glioblastoma multiforme, liver hepatocellular carcinoma, lung adenocarcinoma, and stomach adenocarcinoma (max log2FC = 4.119), with zero expression in all critical somatic organs (max TPM = 0.000). DCAF4L2 has received minimal biological characterization in the published literature, making it a high-priority candidate for experimental validation as a novel retired antigen vaccine target.

#### CT83 (Rank 6)

CT83 (also known as KK-LC-1 or CXorf61) is a cancer-testis antigen originally identified in lung adenocarcinoma that has since been shown to be expressed across multiple common solid cancers including gastric, breast, and lung cancers (Fukuyama *et al*., 2006). CT83 demonstrates strong epigenetic re-activation in tumor tissue, with one of the most negative hypomethylation signals among protein-coding candidates in the final list (delta M-value = -1.572), and is significantly upregulated across 4 cancer types (max log2FC = 4.634) with zero expression in all critical somatic organs (max TPM = 0.000). Despite emerging interest in CT83 as a TCR-based immunotherapy target (Li *et al*., 2022), its potential as a preventive vaccine target within the retired antigen framework has not been explored.

#### TEX19 (Rank 9)

TEX19 (Testis Expressed 19) is a germline-restricted transposon repressor required during spermatogenesis and early embryogenesis for the suppression of retrotransposable elements and maintenance of genomic stability (Ollinger *et al*., 2008). TEX19 has been classified as a cancer-testis gene whose re-expression in tumor tissue is required to maintain cancer cell proliferative potential, with high TEX19 expression linked to poor prognosis in multiple cancer types (Planells-Palop *et al*., 2017). In the RADAR pipeline, TEX19 is epigenetically silenced in normal somatic tissues (normal M-value = 3.38) and is significantly upregulated in breast invasive carcinoma, lung adenocarcinoma, and stomach adenocarcinoma (max log2FC = 4.000, padj = 3.28e-41), representing the most statistically significant tumor re-expression signal in the entire pipeline. TEX19 is targeted by 33 downregulated miRNAs, the highest miRNA regulatory burden among all candidates, providing strong mechanistic evidence for coordinated post-transcriptional de-repression in tumor tissue. It is essentially absent in critical somatic organs (max TPM = 0.030), supporting a favorable autoimmunity safety profile.

### 3.5 Dual Breast and Ovarian Cancer Candidates

Extending the pipeline to TCGA-OV identified 20 candidates shared between breast and ovarian cancer, including MAGEC1, CBX2, KISS1, DRC7, and AACSP1. These represent potential targets for a single vaccine providing protection against both malignancies, consistent with Tuohy’s vision of a combined breast and ovarian cancer vaccine for BRCA1/2 mutation carriers (Mazumder *et al*., 2017; Sakalar *et al*., 2015).

### 3.6 Autoimmunity Safety

The safety screening module eliminated 10 candidates that passed the core discovery layers but showed expression in critical somatic organs above the autoimmunity safety threshold (maximum GTEx TPM > 5) upon cross-referencing with GTEx normal tissue expression data: ZBED2 (7.05), BHLHA15 (28.05), RGS7 (29.10), SHROOM2 (6.95), MMP3 (9.82), ELOVL2 (10.84), TCL1A (7.67), DHRS2 (6.91), BPIFB1 (7.08), and FAM83A-AS1 (5.58). Notably, ZBED2 showed the strongest dual-cancer expression signal in the entire pipeline, upregulated in both BRCA and TCGA-OV tumors, yet exhibited a maximum GTEx TPM of 7.05 in critical somatic organs, disqualifying it as a vaccine candidate due to unacceptable autoimmunity risk. This finding underscores the importance of cross-referencing tumor re-expression signals against normal tissue expression in a second independent dataset, as TCGA tumor upregulation alone was insufficient to identify this safety concern.

### 3.7 Vaccine Strategy Implications

HPA subcellular localization data revealed that most top candidates are intracellular, with primary localization to the nucleoplasm, cytosol, or mitochondria. For these candidates, peptide, mRNA, or viral vector vaccine strategies would be most appropriate, as intracellular proteins are processed and presented on MHC class I molecules to generate cytotoxic T-cell responses (Melief *et al*., 2015). Among the final 20 candidates, those with evidence of surface accessibility or secretion, including COL22A1, CPA6, and LALBA, may additionally be amenable to antibody-based vaccine strategies, as their extracellular localization allows direct antibody recognition without requirement for antigen processing and presentation. This distinction between intracellular and surface-accessible candidates will inform vaccine formulation decisions for experimental validation.

## 4. Discussion

RADAR represents the first systematic computational pipeline specifically designed for retired antigen discovery. Several aspects of the results merit discussion.

The pipeline’s ability to independently validate LALBA and AMH, two targets identified through years of hypothesis-driven wet lab work by Tuohy and colleagues (Tuohy *et al*., 2016; Mazumder *et al*., 2017), demonstrates that the retired antigen concept has a strong computational signature detectable across publicly available databases. It should be noted that while our pipeline detected AMH upregulated in TCGA-OV primary tumors, Tuohy’s ovarian vaccine program targets the extracellular domain of the AMH receptor AMHR2, a distinction that underscores the value of combining computational discovery with mechanistic experimental follow-up. The rediscovery of MAGEA1, MAGEC1, and SSX1 further confirms that the cancer-testis antigen family represents the genomic archetype of retired antigen biology (Simpson *et al*., 2005), and validates the specificity of the tissue-specificity and methylation filters.

The requirement for methylation confirmation in Layer 3 proved critical. DNA methylation is a primary mechanism by which germline-active and reproductive-stage genes are silenced in somatic tissues (Baylin and Jones, 2016; Jones and Baylin, 2002). Genes passing RNA expression filters but lacking a methylation retirement signature are likely regulated by other mechanisms such as transcription factor loss or chromatin remodeling, and may not exhibit the stable, heritable silencing that characterizes true retired antigens. The methylation filter reduced candidates from 1,729 to 84, a 95% reduction, suggesting that only a small fraction of tissue-specific genes achieve their specificity through epigenetic retirement.

The safety screening module eliminated 10 candidates including ZBED2, which had the strongest dual-cancer RNA expression signal in the entire pipeline, based on GTEx critical organ expression exceeding the autoimmunity safety threshold. This result underscores a fundamental principle of retired antigen discovery: tumor re-expression signal strength alone is insufficient to identify safe vaccine candidates, and cross-referencing against normal tissue expression in a second independent dataset is essential before any candidate advances to experimental testing. Future iterations of RADAR should incorporate protein-level quantification from tissue proteomics databases where available to further strengthen safety assessments.

The miRNA regulatory network analysis revealed that most candidates are co-regulated by a shared set of tumor-suppressor miRNAs that are downregulated across multiple cancer types, including hsa-mir-139, hsa-mir-145, hsa-mir-143, and hsa-mir-195. This suggests that retired antigen re-expression in cancer may be mechanistically linked to a shared miRNA network disruption that accompanies carcinogenesis, rather than arising through independent stochastic events in each tumor type (Esquela-Kerscher and Slack, 2006; Garzon *et al*., 2009). This co-regulation has implications for understanding why certain cancer types may be more amenable to retired antigen vaccine strategies.

A notable limitation of RADAR is its dependence on TCGA cohort composition. For cancer types where the TCGA cohort is dominated by a single histological subtype, RADAR may incorrectly exclude retired antigen candidates relevant to minority subtypes that are not well represented in TCGA. Inhibin-alpha (INHA) illustrates this limitation directly. While INHA has been experimentally validated as a therapeutic vaccine target for testicular stromal cell tumors including Leydig and Sertoli cell tumors (Aguilar *et al*., 2017), RADAR fails INHA at Layer 2 because TCGA-TGCT is composed predominantly of germ cell tumors, in which INHA is appropriately downregulated. This false negative result arises not from a flaw in the retired antigen biology of INHA but from the absence of a dedicated testicular stromal cell tumor cohort in TCGA. Future iterations of RADAR should incorporate subtype-stratified expression data and expand to include rare tumor subtypes currently underrepresented in TCGA cohorts.

Several additional limitations should be noted. First, the cross-cancer confirmation module used only 20 TCGA-OV tumor samples due to the limited availability of ovarian cancer expression data with paired normal tissue. Future analyses should incorporate larger ovarian cancer cohorts and use GTEx ovary and fallopian tube expression as the normal tissue comparator. Second, age-stratified GTEx analysis was not implemented in the current pipeline. Incorporating age-stratified expression data would further refine candidates by confirming progressive silencing with age, which is central to the retired antigen hypothesis (Tuohy *et al*., 2020; Tuohy, 2014). Third, single-cell RNA-seq data, which would identify the specific cell types within tumors expressing retired antigens, was not incorporated in the current analysis but should be considered in future iterations to improve cellular resolution of candidate expression patterns.

Finally, the unprognostic designation for most candidates in HPA prognostic data is expected and should not be interpreted as a negative finding. Retired antigens are re-expressed broadly across tumor cells regardless of clinical aggressiveness, which is precisely why they are attractive preventive vaccine targets. They do not require tumor selection for high expression to present a meaningful immune target, and their broad re-expression makes them relevant across the full spectrum of disease rather than only in aggressive subtypes.

## 5. Conclusion

RADAR, implemented on a standard server, provides the first systematic multi-omics computational framework for retired antigen discovery, integrating four sequential core discovery layers with three automated characterization modules. Among 56,200 screened genes derived from GTEx, the pipeline identified 20 high-confidence candidates passing all filters, independently validating known targets including LALBA (Tuohy *et al*., 2016), AMH (Mazumder *et al*., 2017), and established cancer-testis antigens MAGEA1, MAGEC1, and SSX1 (Simpson *et al*., 2005) as positive controls. Among novel protein-coding candidates not previously characterized as retired antigen vaccine targets, DCAF4L2, COX7B2, and TEX19 emerge as the highest priority for experimental validation, demonstrating zero or near-zero expression in critical somatic organs, strong epigenetic silencing in normal tissue confirmed by DNA methylation, and significant re-expression across multiple TCGA cancer types. CT83, while a known cancer-testis antigen, is highlighted as an underexplored vaccine candidate with one of the strongest hypomethylation signals in the dataset.

The pipeline is extensible to any cancer type for which GTEx normal tissue baseline and TCGA multi-omics data are available, and the characterization modules execute automatically as relevant subtype and cross-cancer data become available for a given cancer type. As demonstrated by the INHA analysis, RADAR also correctly excludes candidates that fail the retired antigen criteria, providing a principled and reproducible basis for prioritizing experimental resources. These results support the feasibility of computationally-guided retired antigen discovery as a systematic approach to expanding the cancer immunoprevention pipeline beyond individually characterized targets identified through hypothesis-driven wet lab work (Tuohy, 2014; Tuohy *et al*., 2020; Aguilar *et al*., 2017).

## Data Availability

All pipeline scripts and final candidate tables are publicly available at GitHub: [repository URL to be added upon acceptance]. Primary data sources are publicly available from GTEx (gtexportal.org), TCGA via GDC (portal.gdc.cancer.gov), Human Protein Atlas (proteinatlas.org), and UCSC Xena (xenabrowser.net). miRNA target predictions were obtained from TargetScan v8 (targetscan.org). Processed institutional multi-omics data including pan-cancer differential expression, DNA methylation M-values, and miRNA differential expression parquet files are available from the corresponding author on reasonable request.

## Acknowledgements

The authors gratefully acknowledge the foundational contributions of the late Vincent K. Tuohy, Ph.D. to the retired antigen hypothesis and cancer immunoprevention research. This work was supported by the Western Reserve Academy Cancer Immunology Program. Artificial intelligence tools (Claude, Anthropic) were used to assist with manuscript editing and language refinement. All scientific content, analyses, interpretations, and conclusions are solely the authors’ own.

## References

1. Tuohy, V. K., Johnson, J. M., & Mazumder, S. (2020). Primary immunoprevention of adult onset cancers by vaccinating against retired tissue-specific self-proteins. Seminars in Immunology, 47, 101392. 10.1016/j.smim.2020.101392

2. Jaini, R., Kesaraju, P., Johnson, J. M., Altuntas, C. Z., Jane-Wit, D., & Tuohy, V. K. (2010). An autoimmune-mediated strategy for prophylactic breast cancer vaccination. Nature Medicine, 16(7), 799–803. 10.1038/nm.2161

3. Mazumder, S., Johnson, J. M., Swank, V., Dvorina, N., Martelli, E., Ko, J., & Tuohy, V. K. (2017). Primary immunoprevention of epithelial ovarian carcinoma by vaccination against the extracellular domain of anti-Mullerian hormone receptor II. Cancer Prevention Research, 10(11), 612–624. 10.1158/1940-6207.CAPR-17-0154

4. Sakalar, C., Mazumder, S., Johnson, J. M., Altuntas, C. Z., Jaini, R., Aguilar, R., Naga Prasad, S. V., Connolly, D. C., & Tuohy, V. K. (2015). Regulation of murine ovarian epithelial carcinoma by vaccination against the cytoplasmic domain of anti-Mullerian hormone receptor II. Journal of Immunology Research, 2015, 630287. 10.1155/2015/630287

5. Johnson, J., Budd, G. T., Rhoades, E., & Cleveland Clinic/Anixa Biosciences. (2025). Final results of a Phase I trial of alpha-lactalbumin (aLA) vaccine for breast cancer. Poster presentation at the 2025 San Antonio Breast Cancer Symposium, December 11, 2025, San Antonio, TX. ClinicalTrials.gov: NCT04674306.

6. GTEx Consortium. (2020). The GTEx Consortium atlas of genetic regulatory effects across human tissues. Science, 369(6509), 1318–1330. 10.1126/science.aaz1776

7. Tomczak, K., Czerwinska, P., & Wiznerowicz, M. (2015). The Cancer Genome Atlas (TCGA): an immeasurable source of knowledge. Contemporary Oncology, 19(1A), A68–A77. 10.5114/wo.2014.47136

8. Uhlen, M., Zhang, C., Lee, S., Sjostedt, E., Fagerberg, L., Bidkhori, G., Benfeitas, R., Arif, M., Liu, Z., Edfors, F., Sanli, K., von Feilitzen, K., Oksvold, P., Lundberg, E., Hober, S., Nilsson, P., Mattsson, J., Schwenk, J. M., Brunnstrom, H., Glimelius, B., & Ponten, F. (2017). A pathology atlas of the human cancer transcriptome. Science, 357(6352), eaan2507. 10.1126/science.aan2507

9. Agarwal, V., Bell, G. W., Nam, J. W., & Bartel, D. P. (2015). Predicting effective microRNA target sites in mammalian mRNAs. eLife, 4, e05005. 10.7554/eLife.05005

10. Tuohy, V. K. (2014). Retired self-proteins as vaccine targets for primary immunoprevention of adult-onset cancers. Expert Review of Vaccines, 13(12), 1447–1462. 10.1586/14760584.2014.953063

11. Fan, T., Zhang, M., Yang, J., Zhu, Z., Cao, W., & Dong, C. (2023). Therapeutic cancer vaccines: advancements, challenges, and prospects. Signal Transduction and Targeted Therapy, 8(1), 450. 10.1038/s41392-023-01674-3

12. Xie, N., Shen, G., Gao, W., Huang, Z., Huang, C., & Fu, L. (2023). Neoantigens: promising targets for cancer therapy. Signal Transduction and Targeted Therapy, 8(1), 9. 10.1038/s41392-022-01270-x

13. Du, P., Zhang, X., Huang, C. C., Jafari, N., Kibbe, W. A., Hou, L., & Lin, S. M. (2010). Comparison of Beta-value and M-value methods for quantifying methylation levels by microarray analysis. BMC Bioinformatics, 11, 587. 10.1186/1471-2105-11-587

14. Jones, P. A., & Baylin, S. B. (2002). The fundamental role of epigenetic events in cancer. Nature Reviews Genetics, 3(6), 415–428. 10.1038/nrg816

15. Love, M. I., Huber, W., & Anders, S. (2014). Moderated estimation of fold change and dispersion for RNA-seq data with DESeq2. Genome Biology, 15(12), 550. 10.1186/s13059-014-0550-8

16. Garzon, R., Calin, G. A., & Croce, C. M. (2009). MicroRNAs in cancer. Annual Review of Medicine, 60, 167–179. 10.1146/annurev.med.59.053006.104459

17. Bartel, D. P. (2004). MicroRNAs: genomics, biogenesis, mechanism, and function. Cell, 116(2), 281–297. 10.1016/s0092-8674(04)00045-5

18. Krek, A., Grun, D., Poy, M. N., Wolf, R., Rosenberg, L., Epstein, E. J., MacMenamin, P., da Piedade, I., Gunsalus, K. C., Stoffel, M., & Rajewsky, N. (2005). Combinatorial microRNA target predictions. Nature Genetics, 37(5), 495–500. 10.1038/ng1536

19. Fukuyama, T., Hanagiri, T., Takenoyama, M., Ichiki, Y., Mizukami, M., So, T., Sugaya, M., Sugio, K., & Yasumoto, K. (2006). Identification of a new cancer/germline gene, KK-LC-1, encoding an antigen recognized by autologous CTL induced on human lung adenocarcinoma. Cancer Research, 66(9), 4922–4928. 10.1158/0008-5472.CAN-05-3840

20. Planells-Palop, V., Hazazi, A., Feichtinger, J., Jezkova, J., Thallinger, G., Alsiwiehri, N. O., Almutairi, M., Parry, L., Wakeman, J. A., & McFarlane, R. J. (2017). Human germ/stem cell-specific gene TEX19 influences cancer cell proliferation and cancer prognosis. Molecular Cancer, 16(1), 84. 10.1186/s12943-017-0653-4

21. Li, Q., Hu, W., Liao, B., Song, C., & Li, L. (2022). Natural high-avidity T-cell receptor efficiently mediates regression of cancer/testis antigen 83 positive common solid cancers. Journal for Immunotherapy of Cancer, 10(7), e004713. 10.1136/jitc-2022-004713

22. Ollinger, R., Childs, A. J., Burgess, H. M., Speed, R. M., Lundegaard, P. R., Reynolds, N., Gray, N. K., Cooke, H. J., & Adams, I. R. (2008). Deletion of the pluripotency-associated Tex19.1 gene causes activation of endogenous retroviruses and defective spermatogenesis in mice. PLoS Genetics, 4(9), e1000199. 10.1371/journal.pgen.1000199

23. Dewaker, V., Park, S. T., Jun Lee, J., & Kim, H. S. (2025). Discovery and exploration of small molecule binders for CT83. ACS Omega, 10(22), 22884–22908. 10.1021/acsomega.5c00053

24. Melief, C. J., van Hall, T., Arens, R., Ossendorp, F., & van der Burg, S. H. (2015). Therapeutic cancer vaccines. Journal of Clinical Investigation, 125(9), 3401–3412. 10.1172/JCI80009

25. Aguilar, R., Johnson, J. M., Barrett, P., & Tuohy, V. K. (2017). Vaccination with inhibin-alpha provides effective immunotherapy against testicular stromal cell tumors. Journal for ImmunoTherapy of Cancer, 5(1), 37. 10.1186/s40425-017-0237-2

26. Baylin, S. B., & Jones, P. A. (2016). Epigenetic determinants of cancer. Cold Spring Harbor Perspectives in Biology, 8(9), a019505. 10.1101/cshperspect.a019505

27. Esquela-Kerscher, A., & Slack, F. J. (2006). Oncomirs: microRNAs with a role in cancer. Nature Reviews Cancer, 6(4), 259–269. 10.1038/nrc1840

28. Simpson, A. J., Caballero, O. L., Jungbluth, A., Chen, Y. T., & Old, L. J. (2005). Cancer/testis antigens, gametogenesis and cancer. Nature Reviews Cancer, 5(8), 615–625. 10.1038/nrc1669

29. Tuohy, V. K., Jaini, R., Johnson, J. M., Loya, M. G., Wilk, D., Downs-Kelly, E., & Mazumder, S. (2016). Targeted vaccination against human alpha-lactalbumin for immunotherapy and primary immunoprevention of triple negative breast cancer. Cancers, 8(6), 56. 10.3390/cancers8060056

30. Chen, Y. X., Chen, H., Rong, Y., Jiang, F., Chen, J. B., Duan, Y. Y., & He, J. (2020). An integrative multi-omics network-based approach identifies key regulators for breast cancer. Computational and Structural Biotechnology Journal, 18, 2826–2835. 10.1016/j.csbj.2020.10.001

31. Jones, P. A., & Baylin, S. B. (2002). The fundamental role of epigenetic events in cancer. Nature Reviews Genetics, 3(6), 415–428. 10.1038/nrg816

